# Brain-Regional Gene Expression Imputed from the Blood Transcriptome by *BrainGENIE* Recapitulates Dysregulation Observed in the Postmortem Brain in Alzheimer’s Disease

**DOI:** 10.1101/2025.10.13.25337831

**Authors:** Jiahui Hou, Ali Razavi, Shu-Ju Lin, Chunling Zhang, William S. Kremen, Christine Fennema-Notestine, Jeremy Elman, Peter Holmans, Stephen V. Faraone, Chris Gaiteri, Jonathan L. Hess, Stephen J. Glatt

**Author notes:** Jiahui Hou and Ali Razavi contributed equally to this work and are co-first authors. **Please address correspondence to:** Stephen J. Glatt, Ph.D., SUNY Upstate Medical University, 750 East Adams St., Syracuse, NY 13210 U.S.A., 1.

## Abstract

Studying brain gene expression in Alzheimer’s Disease (**AD**) presents significant challenges as postmortem brain tissue data is difficult to access, cannot be used to guide donor treatment, may be affected by confounding environmental factors before and after death and is difficult to link to early AD states or disease progression. To circumvent these limitations, several studies have tested blood transcriptome biomarkers for AD. However, gene-expression levels in the blood have limited correlation with those in the brain. Therefore, to test the potential of monitoring Alzheimer’s progression with peripheral data, we used a transcriptome-imputation method, to identify brain-region-specific AD-associated gene-expression differences in multiple cohorts with available blood-based transcriptome data. This approach provides a high-resolution image of the AD-associated molecular differences in the brains of affected individuals *actively living with disease*. We analyzed eight AD studies (777 AD cases, 779 cognitively unimpaired controls) in which we imputed brain-regional gene expression in 10 brain areas, using Brain Gene Expression and Network Imputation Engine (*BrainGENIE)*. Hundreds of differentially expressed genes (**DEG**s) associated with AD were identified in nine brain regions, with anterior cingulate cortex and amygdala showing the most differential expression. AD-associated genes were enriched in pathways related to proteostasis, mitochondrial dysfunction, and immune activation, among others. We observed significant congruence between imputed AD-associated changes and those directly measured in the dorsolateral prefrontal cortex and cerebellum. These transcriptomic changes can be leveraged in future *in vitro* studies focused on pathogenesis, or as the targets of novel therapeutic developments. In conclusion, we demonstrated the scope and utility of brain expression imputation from the peripheral transcriptome, laying the groundwork for biomarker discovery and prospective studies on the aging brain and AD.

## Introduction

There are over one hundred different hypotheses for the pathogenesis of AD (Bermejo-Pareja and Del Ser, 2024) and mixed pathologies are normal to observe in the aged brain (Bai et al., 2014). Long-standing hypotheses focus on the effects of amyloid and intracellular tau (Leandro et al., 2018)(Shigemizu et al., 2021), while recent brain omics studies have indicated the relevance of additional molecular systems, including neuroinflammation, neurovascular dysfunction, cholinergic deficit, and calcium dyshomeostasis (Alzheimer’s Association, 2025; Bermejo-Pareja and Del Ser, 2024). Brain omic measurements may be useful in determining the overall impact of complex AD genetics and multiple comorbid pathologies. Over the course of decades, the largest brain omic studies have grown to include several hundred brains (Samsuddin et al., 2018)(Mathys et al., 2021) and help to describe convergent effects onto particular molecular mechanisms. However, brain omics are challenging to obtain and rarely found for all genetic backgrounds. Therefore, peripheral omics offer the potential to make such brain studies more accessible, increase our ability to define samples from the earliest stages of AD and permit more rapid therapeutic assessments – if their own limitations can be addressed.

Serially sampled peripheral gene expression may have select advantages over brain gene expression, because it can be acquired around the time of the first disease symptoms, which is likely a key period in which to identify early and influential drivers of disease. While brain expression can certainly also capture this period, the timing of its single snapshot can occur at any disease stage, so such early onset data is generated essentially at random. Furthermore, from single timepoint data, it may be difficult to determine if a particular cognitive test represents the beginning of disease in light of the instability of cognitive measures (Lunnon et al., 2013). There are also strong statistical advantages to capturing AD from a blood perspective, because comparisons are made within-person, at different time points in diseases. In such comparisons each subject serves as their own control, limiting noise from real biological individuality that is not related to disease. Peripheral studies may more broadly and accurately represent disease, because the largest brain omics studies are predominantly in highly educated Caucasian populations. While there are new cohorts that represent broader genetic backgrounds (Hou et al., 2024), their sample size is much smaller, as brain donation is a practical barrier to participation. Thus, there is at least theoretical potential for peripheral transcriptome studies to supply critically timed and broadly-based information on AD etiology, provided that they are sufficiently reflective of central pathology.

While blood-based biomarkers and diagnostics for Alzheimer’s have appeal as accessible proxies, they must confront more limited relationships to at least some disease processes. Attempting to find disease modulators against peripheral amyloid and tau markers through GWAS has found a small number of loci, all likely highly related to amyloid, tau or APOE (Wan et al., 2020) (Duran-Aniotz et al., 2022). While the observability of genetic signals related to peripheral disease markers bodes well for disease representation, they were all closely tied to the marker itself, as opposed to showing far-flung interacting factors. While brain transcriptome studies have shown hundreds of disease-correlated transcripts, the ability of blood transcriptome to represent disease may be limited due to small correlations between the brain and blood (Tylee et al., 2013; Sullivan et al., 2006), potentially due to genetic, epigenetic, or other regulatory effects shared across tissues (GTEx Consortium, 2020; GTEx Consortium, 2015). Blood transcription studies in Alzheimer’s on brain molecules for the transcriptomic state and activity of the brain (Lu et al., 2021; Xue et al., 2020; Ji et al., 2022; Neshan et al., 2020; Leandro et al., 2018). These studies found enrichment for immune and synaptic systems, findings which are certainly consonant with brain transcriptome studies, but overall, not highly consistent with each other, and the samples sizes are small (n<40 per class) in relation to current brain-based studies. While findings are not negative, it is not clear to what extent they capture disease processes.

Recent advances in brain-transcriptome imputation have significantly increased the number of gene transcripts whose abundance can be estimated in the brain, with models that go beyond one-to-one correlations of a given molecule across tissue (Hess et al., 2023). The Brain Gene Expression and Network Imputation Engine (*BrainGENIE*) capitalizes on robust principal components (PCs) of gene expression in peripheral blood, which represent the activity across multiple transcripts and pathways, using those to impute expression of individual transcripts in specific brain regions. These predictions include transcripts that are solely expressed in the brain - completely absent in blood. Unlike genetic-based imputation methods (*e.g.*, *PrediXcan (Gamazon et al., 2015)*, *FUSION (Gusev et al., 2016)*, *EpiXcan (Hou et al., 2024)*), *BrainGENIE* has the potential to account for both genetic and non-genetic factors influencing gene-expression profiles, not only capturing well-established disease-related changes in gene expression measured directly in the brain, and also novel disease signatures in the brains of living individuals. Indeed, *BrainGENIE* outperforms *PrediXcan* in recapitulating disorder-related changes in autism, bipolar disorder and schizophrenia (Hess et al., 2023). The increased scope of predictions from this method may be useful in representing peripheral effects of Alzheimer’s brain processes.

To provide an atlas of brain-regional transcriptomic signatures associated with AD in living individuals, which may shed light on mechanisms relevant to AD pathology, we identify genes imputed from blood transcriptome PCs that are differentially expressed within the brain in individuals living with AD compared to cognitively unimpaired participants (CU). We demonstrate the possibility of longitudinal assessments in older adults as they pass through periods of AD risk, progression, and treatment by using *BrainGENIE* to impute brain data. Results are validated in comparison to AD-related changes in brain gene expression from previous *postmortem* brain studies. The effect of these results is to determine the extent and strength of imputed relationships, to inform future *in silico, in vitro* and *in vivo* projects by significantly increasing the available AD-related brain data and providing novel molecular targets for preclinical and clinical studies.

## Materials and Methods

### Peripheral-blood transcriptomic studies of AD

We included eight peripheral-blood gene-expression studies with 777 AD cases and 779 CU controls from ADNI, Bai *et al*. (2014), Leandro *et al*. (2018), AddNeuroMed batch-1, AddNeuroMed batch-2, Religious Orders Study/Memory and Aging Project (ROSMAP) batch-1/2, Samsudin *et al*. (2016), and Shigemizu *et al*. (2020) (**Table 1**), which represents all large AD peripheral studies. The number of protein-coding genes included in the analysis ranged from 9,951 to 18,168 across the eight studies. As the datasets in our analysis were from previous studies with various sampling and profiling procedures, data were reprocessed using a consistent set of tools and a consistent pipeline to mitigate unwanted between-study variance that could obfuscate biologically meaningful signals. Our previous transcriptomic meta-analysis [Hou, 2024] reported these data-processing procedures (*i.e*., gene-expression quality control, sample outlier detection, normalization, and batch-effect correction) (Hou et al., 2024). Specifically, quantile-normalization was applied to all microarray studies. RNA-sequencing data were normalized to effective library size by the trimmed mean of M values (Hou et al., 2024). Table 1 summarizes the demographic characteristics of the participants in each study.

**Table 1.** Characteristics of donors to the Eight Alzheimer’s disease blood transcriptomic studies.

| Study | Diagnosis | Mean(SD) Age | Age Range | Percent Female | # of Cases | Sequencing Platform | # of Genes Detected |
| --- | --- | --- | --- | --- | --- | --- | --- |
| Lovestone_1 | AD | 75.60 (6.86) | 58 - 88 | 71.88 | 96 | Illumina HumanHT-12 v3 Expression BeadChip | 16456 |
| Lovestone_1 | CTL | 72.63 (6.72) | 52 - 87 | 58.49 | 106 | Illumina HumanHT-12 v3 Expression BeadChip | 16456 |
| Lovestone_2 | AD | 77.50 (6.70) | 60 - 90 | 63.25 | 117 | Illumina HumanHT-12 v3 Expression BeadChip | 17775 |
| Lovestone_2 | CTL | 75.70 (5.99) | 63 - 89 | 61.54 | 130 | Illumina HumanHT-12 v3 Expression BeadChip | 17775 |
| ROSMAP_b1b2 | AD | 86.23 (4.69) | 67.76454 - 90 | 65.45 | 55 | Affymetrix Human Genome U133 Plus 2.0 | 14638 |
| ROSMAP_b1b2 | CTL | 85.28 (5.49) | 70.78987 - 90 | 66.67 | 87 | Affymetrix Human Genome U133 Plus 2.0 | 14638 |
| Samsudin16 | AD | 77.68 (5.81) | 64 - 94 | 50 | 90 | Illumina HumanHT-12 v4 Expression BeadChip | 18009 |
| Samsudin16 | CTL | 75.23 (7.17) | 65 - 94 | 47.78 | 90 | Illumina HumanHT-12 v4 Expression BeadChip | 18009 |
| Shigemizu20 | AD | 79.55 (5.85) | 65 - 93 | 68.4 | 269 | Illumina HiSeq 2000 RNA-Seq | 9952 |
| Shigemizu20 | CTL | 71.29 (5.07) | 60 - 83 | 45.05 | 91 | Illumina HiSeq 2000 RNA-Seq | 9952 |
| Leandr18 | AD | 79.36 (6.56) | 65 - 91 | 63.64 | 22 | Agilent 4x44K Whole Human Genome | 13797 |
| Leandr18 | CTL | 77.31 (6.17) | 69 - 85 | 76.92 | 13 | Agilent 4x44K Whole Human Genome | 13797 |
| Bai14 | AD | 75.94 (6.73) | 60 - 86 | 70.59 | 17 | Agilent 4x44K Whole Human Genome | 12253 |
| Bai14 | CTL | 73.71 (4.08) | 68 - 82 | 66.67 | 21 | Agilent 4x44K Whole Human Genome | 12253 |
| Adni | AD | 77.46 (7.66) | 56 - 92 | 35.34 | 116 | Illumina HumanHT-12 v4 Expression BeadChip | 18171 |
| Adni | CTL | 76.37 (6.51) | 56 - 94 | 52.44 | 246 | Illumina HumanHT-12 v4 Expression BeadChip | 18171 |

### Brain gene-expression imputation using BrainGENIE

We used *BrainGENIE* (Hess et al., 2023) to impute brain-regional gene-expression profiles for 10 different brain regions in the eight above-mentioned blood gene-expression studies of AD. The 10 brain regions included: amygdala, anterior cingulate cortex (ACC; Brodmann area [BA] 24), cerebellum (primarily gray matter), dorsolateral prefrontal cortex (DLPFC, BA9), hippocampus, hypothalamus, caudate nucleus, nucleus accumbens, putamen, and substantia nigra. As described by Hess *et al*. (Hess et al., 2023), *BrainGENIE* was trained on fresh-frozen samples from deceased donors in the Genotype-Tissue Expression (GTEx) project, version 8 (v8). For each AD study, *BrainGENIE* models were trained using the genes detected in the blood of study participants and the blood of donors from GTEx v8. *BrainGENIE* models were trained on the subset of genes jointly found in the blood in the to-be-imputed data and the GTEx V8 data. The algorithm then utilized the subset of genes and samples from the GTEx V8 database to develop imputation models for all brain transcripts present in the GTEx V8 database. The models resulting from this algorithm have high consistency in prediction as demonstrated in our original paper, replicating disease gene-expression pathways in all 10 regions better than all preceding gene-expression imputation models (Hess et al., 2023). Imputation performance was estimated by averaging the per-gene imputation accuracy (Pearson’s correlation, *r*, of observed *vs*. imputed gene-expression) across five independent validation-folds from cross-validation. Genes found to be significantly predicted based on cross-validation in GTEx–defined as a mean cross-validation *R*^2^ ≥ 0.01 and Benjamini–Hochberg false-discovery rate (FDR)-adjusted *p*-value < 0.05–were retained for imputation. A total of 80 brain-imputed datasets (eight AD blood studies * 10 brain regions) were generated using this approach.

### Meta-analysis of genes differentially expressed in AD

After grouping samples based on study and brain region, we used linear regression models to estimate mean differences in imputed gene-expression levels between AD cases and CU comparison participants. We used the *lmFit* function from the *R* package *limma* (Ritchie et al., 2015), which estimates empirical Bayes-moderated *t*-statistics. Our linear regression models specified imputed brain gene expression as the dependent variable and AD diagnosis as the independent variable. We included age and sex as covariates for each analysis. We included self-reported race and RNA integrity number (RIN) as covariates, when available. We applied a data-driven dconvolution approach in the *R* package *dtangle* (Hunt et al., 2019) on the peripheral blood gene-expression profiles to estimate the relative abundance of plasma and seven blood cell-types (*i.e.*, B cell, T cell, dendritic cell, monocytes, mast cell, natural killer cell, and granulocytes). Similarly, we applied the same approach on each imputed brain dataset to estimate the relative abundance of four brain cell-types (neuron, oligodendrocyte, microglia, astrocyte). The estimated proportions of seven blood cell-types and three brain cell-types were used as covariates in each differential gene-expression analysis (We excluded the estimated proportions of plasma and one of the four brain cell-types to prevent multicollinearity.)

To account for hidden confounding effects, we performed a surrogate variable analysis (SVA) (Leek et al., 2012) using the *R* package *SVA* on each blood dataset to identify unmeasured sources of potentially confounding variation called “surrogate variables” (SVs). We detected the SVs after accounting for variation in gene expression explained by AD diagnosis and other known covariates as the adjustment variables (*i.e.*, age, sex, blood-cell proportions, race), and RNA integrity number). Only one blood study (*i.e*., Leandro *et al*., 2018) had identifiable SVs that explained unmodeled variation in gene expression at the default *p* < 0.1. To reduce the loss of degrees of freedom and prevent overfitting in the models, we used the top four detected SVs (from a total of 6) as covariates for the imputed brain datasets from Leandro *et al*., 2018. **Supplementary Table 1** documents the covariates included for each study in the differential gene-expression analysis.

In each brain region, we retained all protein-coding genes present in at least two studies for the meta-analysis. We used a random-effects inverse-variance weighted meta-analysis model to pool differential-expression coefficient estimates and sampling variances of the coefficient estimates per gene across studies per brain region. We chose this model because it accounts for between-study variances and produces conservative estimates of significance in the possible presence of heterogeneity of coefficient estimates. We used the *R* package *metafor* (Viechtbauer, 2010) to perform the random-effects meta-analysis with the DerSimonian-Laird estimator.

### Gene-set over-representation analysis

In each brain region, the *R* package *piano* (Varemo et al., 2013) was used for gene-set enrichment analysis using summary statistics obtained from our differential gene-expression meta-analyses of protein-coding genes. Effect sizes and *p*-values of differential expression were supplied to the method *runGSA*, which computed the gene-set-level statistics across *Reactome* pathways. Two enrichment tests were performed: 1) to test whether gene-sets were enriched for genes up-regulated in AD cases; and 2) to test whether gene-sets were enriched for genes down-regulated in AD cases. In each directional enrichment test, the gene-set-level statistic was derived from the mean of transformed *p*-values. For the upregulation (downregulation) test, the raw *p*-values of the up (down)-regulated genes were transformed (*i.e.*, *p*/2) to range from 0 to 0.5, and the raw *p*-values of the down (up)-regulated genes were transformed (*i.e.*, 1-*p*/2) to range between 0.5 and 1. To reduce the number of broad biological categories tested and to provide better insight into specific underlying biological processes and molecular functions, gene-sets were limited to those with a range of 10 to 200 genes. To determine the significance of the enrichment test, gene labels were permuted 100,000 times and mean differential-expression statistics were recomputed for each permuted gene-set. We used the Bonferroni procedure for multiple-testing correction of the significance values from our gene-set enrichment analyses with a significance value of 0.05.

### Concordance of disease signals between imputed and directly measured gene-expression profiles

We previously meta-analyzed 24 *postmortem* brain studies spanning six brain regions, including three regions for which *BrainGENIE* models can impute transcriptome profiles (cerebellum, DLPFC, and hippocampus) (Hou et al., 2024). The details of the pre-processing of the datasets have been covered in the Hou *et al*. meta-analysis. We sought to determine the comparability of AD-related changes in brain-imputed blood-based gene-expression from *BrainGENIE* and transcriptomic changes measured in the *postmortem* AD brain. To address this we measured the concordance between AD-related differential gene expression (DGE) changes found in brain-imputed measures and DGE changes from directly measured *postmortem* brain gene-expression.

We evaluated how well *BrainGENIE*-imputed brain gene-expression data captured AD-associated changes identified in three *postmortem* brain regions (*i.e*., cerebellum, DLPFC, and hippocampus). We used Spearman’s correlation to measure the concordance of per-gene differential-expression effect sizes across the whole transcriptome between imputed data of 10 brain regions and directly measured *postmortem* brain data from three regions. We used (1 - Spearman’s *rho*) to measure the dissimilarity between any two of the 14 imputed and measured datasets and applied hierarchical clustering to group these studies into distinct clusters. In addition, for each of the three brain regions with both imputed and directly measured gene-expression profiles (*i.e*., cerebellum, DLPFC, and hippocampus), we examined whether the differentially expressed genes (DEGs) found in directly measured data could be recapitulated in the imputed brain expression data from living individuals. Similarly, we evaluated the overlap of AD-associated *Reactome* pathways between imputed and directly measured gene-expression profiles for hippocampus, cerebellum, and DLPFC.

### Single-cell gene ontology analyses

We performed a *post-hoc* analysis of the AD DEGs found in our study to evaluate enrichment of marker genes for cell types found in the adult brain. Normalized single-cell RNA-seq data were obtained from the *BrainSCOPE* dataset from the *PsychENCODE* project (Emani et al., 2024), containing average per-gene expression levels for 17,158 common genes across 24 different brain cell types. Using the package *dtangle* (Hunt et al., 2019), we identified cell-type-specific marker genes in this dataset using the “diff” method, which nominates cell-specific marker genes based on median differences in gene-expression levels. We selected the top 10, 50, 250 and 500 marker genes for each cell type to test for enrichment of AD-associated DEGs with one-tailed Fisher’s exact tests. We adjusted for multiple comparisons using the FDR procedure with a significant *p*-value threshold of 0.05.

### Sources of intra-regional heterogeneity of measured gene-expression

Given diverging results from our correlation analyses comparing imputed and measured data across different brain regions, we performed exploratory *post-hoc* analyses of the measured *postmortem* brain data to further understand the source of the variation in the results. Measured gene-expression levels from the *postmortem* hippocampus, cerebellum, and DLPFC were obtained and processed as described in the meta-analysis (Hou et al., 2024). The pairwise Pearson’s correlation per-transcript between the individual studies for each brain region were calculated. Furthermore, all studies were pooled by the common genes and PCA was performed to examine the heterogeneity of data in each region and between regions. This process was repeated with only the subsets of studies that included RNAseq data. Additionally, we inspected the gene-level Cochran’s Q statistics from our meta-analysis to check for potential interstudy differences with respect to differential expression estimates for AD in the hippocampus, DLPFC, and cerebellum (FDR < 0.05 designated as significant).

To normalize for tissue-extraction location, transcriptome data from *postmortem* hippocampus studies were supplied to a model developed by *Vogel et al.* to examine sub-tissue location of data extraction (Vogel et al., 2020). Utilizing data reported from the Allen Brain Atlas, the common genes between the studies of Miller *et al*., 2008 and Blalock *et al*., 2004 were used to develop two separate models that predict hippocampus data-extraction location. The models were not able to accurately predict the location of extraction in the two studies and thus were excluded from downstream analyses.

## Results

### DEGs in BrainGENIE-imputed Brain Transcriptomes

**Supplementary Table 3** highlights the number of imputed genes in each of the 10 brain regions in the eight studies (ranging from an average of 1,928 in substantia nigra to 8,707 in DLPFC). We retained genes that were present in at least two studies. After combining all studies for each brain region, the number of unique genes in each meta-analysis ranged from 2,195 in substantia nigra to 9,238 in caudate. We identified genes that were significantly differentially expressed (FDR-*p* < 0.05) in AD cases compared with CU comparisons in nine brain regions (all except for substantia nigra), and the number of DEGs ranged from 6 to 705 (**Figure 1A**). **Supplementary Table 4** presents the summary statistics of the significant DEGs (FDR-*p* < 0.05) across the nine brain regions. The largest number of DEGs was found in ACC (*k*_DEGs_ = 705, 17.3% of total imputed genes in region), followed by amygdala (*k*_DEGs_ = 687, 11.9% of total imputed genes in region), and caudate (*k*_DEGs_ = 52, 0.56% of total imputed genes in region). The smallest number of DEGs was identified in cerebellum (*k*_DEGs_ = 6, 0.07% of total imputed genes in region).

**Figure 1.**
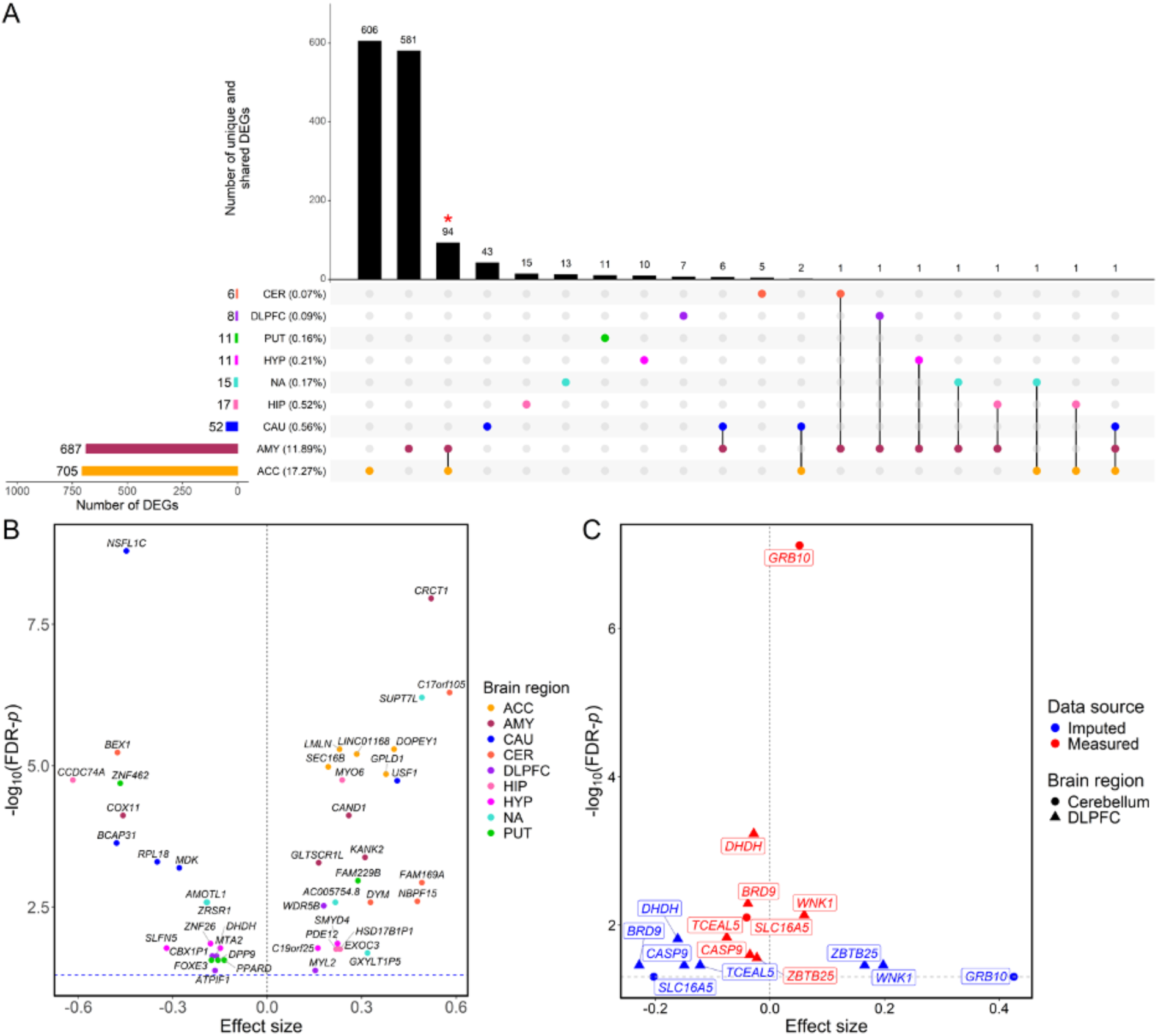
Differentially expressed genes (DEGs) across 10 imputed brain regions. In panel A, the upset plot shows the number of genes that were significantly dysregulated in AD cases compared with CU comparisons (FDR-*p* < 0.05) in each of the 10 imputed brain regions, the number of unique DEGs in a brain region, and the number of shared DEGs across brain regions. The red asterisk denotes cases where the number of DEGs common between that label and DEGs found in measured data is higher than expected by chance with a *p*-value of 0.01. Panel B shows the volcano plot of the top five most significant DEGs in each brain region. Panel C shows the DEGs identified in directly measured *postmortem* cerebellum and DLPFC data that were significantly replicated in the imputed cerebellum and DLPFC data, respectively. For panels B and C, the x-axis shows the effect size of gene-expression differences between AD cases and CU comparisons in each meta-analysis. The y-axis indicates the transformed FDR-corrected *p*-value of the effect-size estimate. A blue horizontal dotted line denotes FDR-*p* = 0.05. Abbreviations: DEGs: differentially expressed genes; AD: Alzheimer’s disease; CU: cognitively unimpaired; FDR: false discovery rate; ACC: anterior cingulate cortex; AMY: amygdala; CAU: caudate; CER: cerebellum; DLPFC: dorsolateral prefrontal cortex; HIP: hippocampus; HYP: hypothalamus; NA: nucleus accumbens; PUT: putamen; SN: substantia nigra.

To evaluate whether gene predictability systematically influenced our differential expression results, we analyzed the relationship between cross-validated prediction performance (Pearson’s r) in GTEx and the magnitude of AD-associated differential expression (absolute t-statistic) in the imputed frontal cortex (BA9). We observed a weak and significant correlation (Spearman’s ρ = 0.056, p = 3.2 × 10⁻⁷).

Most DEGs were region-specific. We evaluated whether pairwise DEGs across the ten brain regions exceeded what would be expected by chance. Only the ACC and the amygdala overlap with 94 different DEGs reaching statistical significance (Figure 1A; Fisher’s exact test, Bonf-p = 6.3×10^-22^).). No other region-pair shared a DEG set that was significant at the multiple-testing threshold.

Because the imputed data for all brain regions were derived from the same sample of blood gene expression data, if a greater number of genes could be reliably imputed in a region, there is, *a priori*, a higher chance that more genes will be found to be dysregulated in that region. However, the correlation between the total number of unique imputable genes in the meta-analysis and the number of DEGs across the 10 brain regions was not significant (Pearson’s *r* = −0.264, *p* = 0.46). To standardize comparisons across different brain regions, we examined the rate of DEGs, which adjusts for the number of unique genes in the meta-analysis. The rate of DEGs—the number of DEGs divided by the number of unique genes in the meta-analysis—ranged from 0.07% to 17.27% across nine brain regions (**Figure 1A**). DEG prevalence significantly differed across regions (χ² = 6 878.6, *df* = 8, *p* < 2 × 10⁻¹⁶). After adjusting for the total number of unique genes included in the meta-analysis, the top three regions with the highest rate of DEGs were still the ACC (17.27%), amygdala (11.89%), and caudate (0.56%).

### Disease Signal Concordance Between Imputed and Directly Measured Gene-expression Profiles

For each of the three brain regions (*i.e*., cerebellum, DLPFC, and hippocampus) that had both imputed and directly measured gene-expression profiles, we examined if the imputed gene-expression data replicated the DEG signals found in the directly measured gene-expression datasets. Among the DEGs (FDR-*p* < 0.05) identified in the directly measured datasets, there were 881, 2,042, and three genes that were present in the current analysis of imputed data in cerebellum, DLPFC and hippocampus, respectively. We found two genes in cerebellum and six genes in DLPFC that were significantly replicated in the imputed data (FDR-*p* < 0.05; **Figure 1C**). The number of significant genes overlapping between imputed and predicted data in the DLPFC was higher than expected by chance (*p* = 0.006). Among the eight significantly replicated genes, seven showed dysregulation in the same direction in both imputed and directly measured data, including upregulated *GRB10* (Growth Factor Receptor Bound Protein 10) and downregulated *SLC16A5* (Solute Carrier Family 16 Member 5) in cerebellum, and downregulated *DHDH* (Dihydrodiol Dehydrogenase), *BRD9* (Bromodomain Containing 9), *CASP9* (Caspase 9), *TCEAL5* (Transcription Elongation Factor A Like 5) and upregulated *WNK1* (WNK Lysine Deficient Protein Kinase 1) in DLPFC (**Figure 1C**). Although fewer than 1% of the individually significant postmortem DEGs also reached FDR in the imputed data, the transcriptome-wide effect sizes between imputed brain gene expression in living participants and measured gene expression from postmortem brain samples were still moderately correlated (ρ = 0.35 for cerebellum, ρ = 0.28 for DLPFC; Fig. 2)

**Figure 2.**
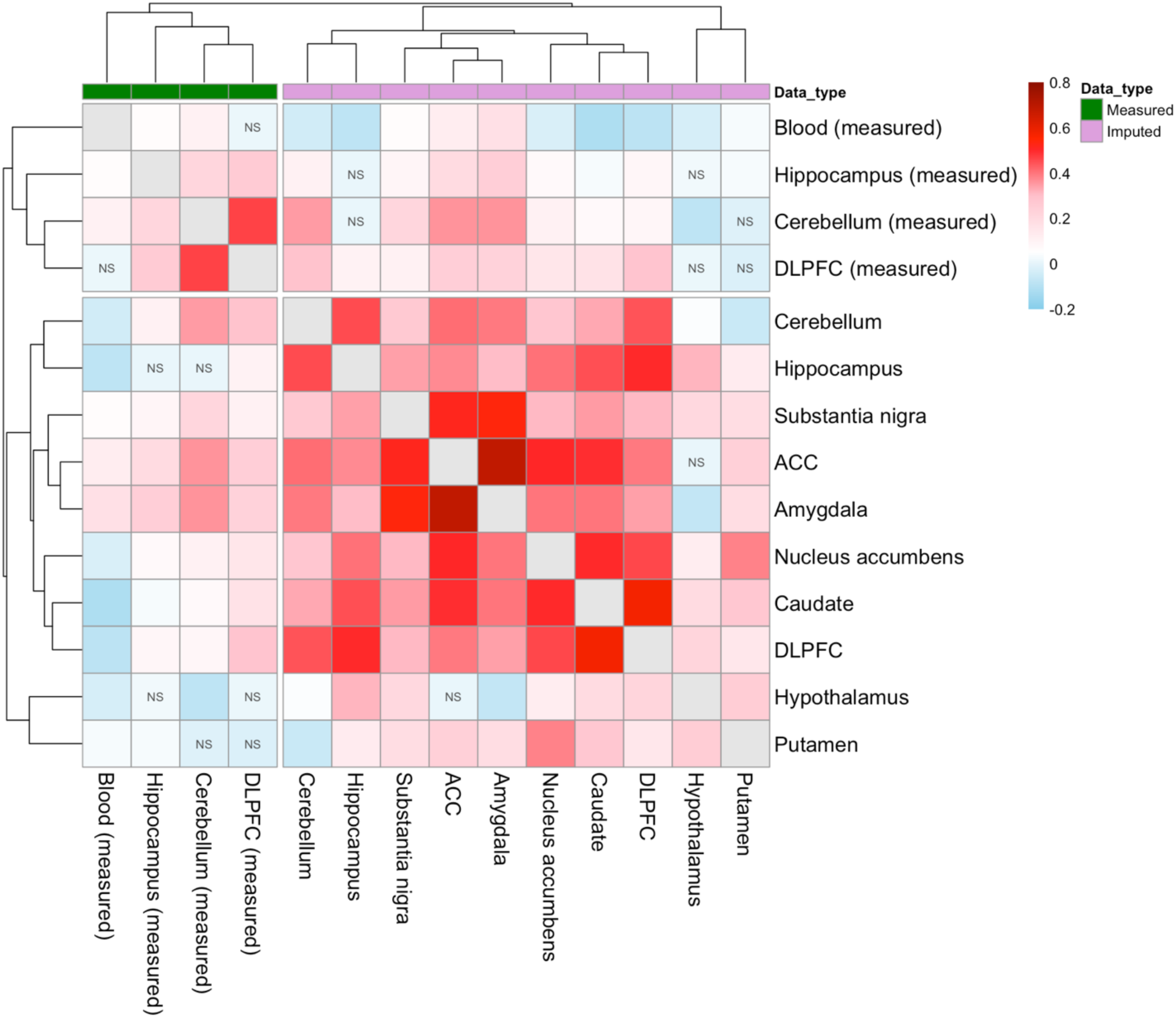
Heatmap of per-gene effect size correlation between directly measured and imputed data. The color of heatmap cells indicates the Spearman’s *rho* of gene differential expression effect size across the whole transcriptome between any two datasets. Red color indicates Spearman’s *rho* > 0 and blue color indicates Spearman’s *rho* < 0. “NS” label in the heatmap cell indicates the correlation was not significant whereas the absence of “NS” indicates significance (*p* < 0.05). The color bar on the top of the heatmaps shows whether the data were directly measured or imputed using *BrainGENIE*. Abbreviations: ACC: anterior cingulate cortex; DLPFC: dorsolateral prefrontal cortex.

### Correlations Between AD-associated DEG Effects Between Regions

We found significant correlations of AD-associated differential-expression effects across the whole transcriptome between all pairs of the 10 imputed brain regions, except for between ACC and hypothalamus. Across imputed brain regions with significant DGE concordance, Spearman’s rho ranged from −0.069 (*p* = 7.8×10^-4^) between amygdala and hypothalamus to 0.687 (*p* = 2.0×10^-283^) between amygdala and ACC (**Figure 2**; **Supplementary Table 2**). We identified two distinct clusters based on the DGE agreement patterns, one comprising directly measured brain data and the other consisting of the imputed data from 10 brain regions (**Figure 2**).

For cerebellum and DLPFC, DGE effect sizes estimated from imputed gene-expression profiles were significantly correlated with those estimated from directly measured gene-expression profiles (Spearman’s ρ = 0.351, *p* = 3.5×10^-180^; Spearman’s ρ = 0.281, *p* = 1.5×10^-144^; respectively). However, we did not find a significant correlation of DGE effects between imputed and directly measured gene-expression data in the hippocampus (Spearman’s ρ = 0.005, *p* = 0.79). Across the three brain regions with both imputed and directly measured postmortem brain expression data (*i.e*., cerebellum, DLPFC, and hippocampus), the correlations of DGE effect sizes between imputed datasets were not significantly different from those between directly measured gene-expression profiles (z-test = 1.96, *p* = 0.052). In other words, inter-region similarity of DGE patterns was not significantly different in imputed vs. measured data.

### Intra-Regional Heterogeneity in Postmortem Brain Studies

The lower correlation observed between imputed and measured hippocampus data, relative to the DLPFC and cerebellum, led to exploratory analyses into the three measured datasets. We evaluated inter-study gene-expression variation within each brain region as a potential contributor to differences in correlations between imputed and measured brain data. We found that the intra-region Pearson’s correlations capturing gene-expression similarity between postmortem hippocampal studies ranged from 0.74 – 0.92, on par with inter-study correlations observed for postmortem DLPFC studies (0.70 – 0.99) and postmortem cerebellum studies (0.58 – 0.94). Further, we assessed inter-study heterogeneity in our gene-level differential expression meta-analyses from postmortem brain datasets by comparing DLPFC, cerebellum, and hippocampus using Cochran’s Q-statistics, which revealed no significant gene-wise difference between studies (*p*-values > 0.05) Lastly, we evaluated a hippocampal substructure prediction model to assess whether dissection differences across postmortem hippocampal studies affected our correlation results. The PCR-LR model from *Vogel et al*, which was trained to predict substructure of the hippocampus based on the continuous variation in gene expression along its spatial gradient, was unable to predict the “ground truth” labels for donors from Blalock *et al*. (CA1) and Miller *et al*. (CA1 and CA3) (AUC < 0.5), and thus could not be used to explain potential heterogeneity in our study.

### Gene-sets Over-represented by AD-associated Genes

For each of the 10 imputed brain regions, we used the meta-analysis summary statistics of the whole transcriptome for gene-set over-representation analysis. Across 10 imputed brain regions, we found a combined total of 546 significantly over-represented Reactome pathways that were distinctly up-regulated in AD cases compared with CU comparisons, which included 327 unique Reactome pathways (FDR-p < 0.05; **Supplementary Table 5**). In addition, we identified a total of 402 Reactome pathways, including 185 unique pathways, that were significantly over-represented and distinctly down-regulated in AD cases (FDR-p < 0.05; **Supplementary Table 6**). The number of significantly up-regulated and down-regulated *Reactome* pathways in each brain region is shown in **Figure 3A-3B**. All the significantly enriched Reactome pathways in the hippocampus were up-regulated, and only down-regulated Reactome pathways were identified in putamen. In ACC and DLPFC, down-regulated pathways significantly outnumbered up-regulated pathways (one-tailed binomial p = **3.13×10⁻³⁰** and **2.03×10⁻⁵**, respectively). The amygdala also showed a down-regulation predominance (p = **3.08×10⁻¹⁴**). In contrast, up-regulated pathways predominated in hypothalamus and nucleus accumbens (p = **3.09×10⁻⁴¹** and **5.65×10⁻⁹**, respectively), while caudate, cerebellum, and substantia nigra showed no directional dominance

**Figure 3.**
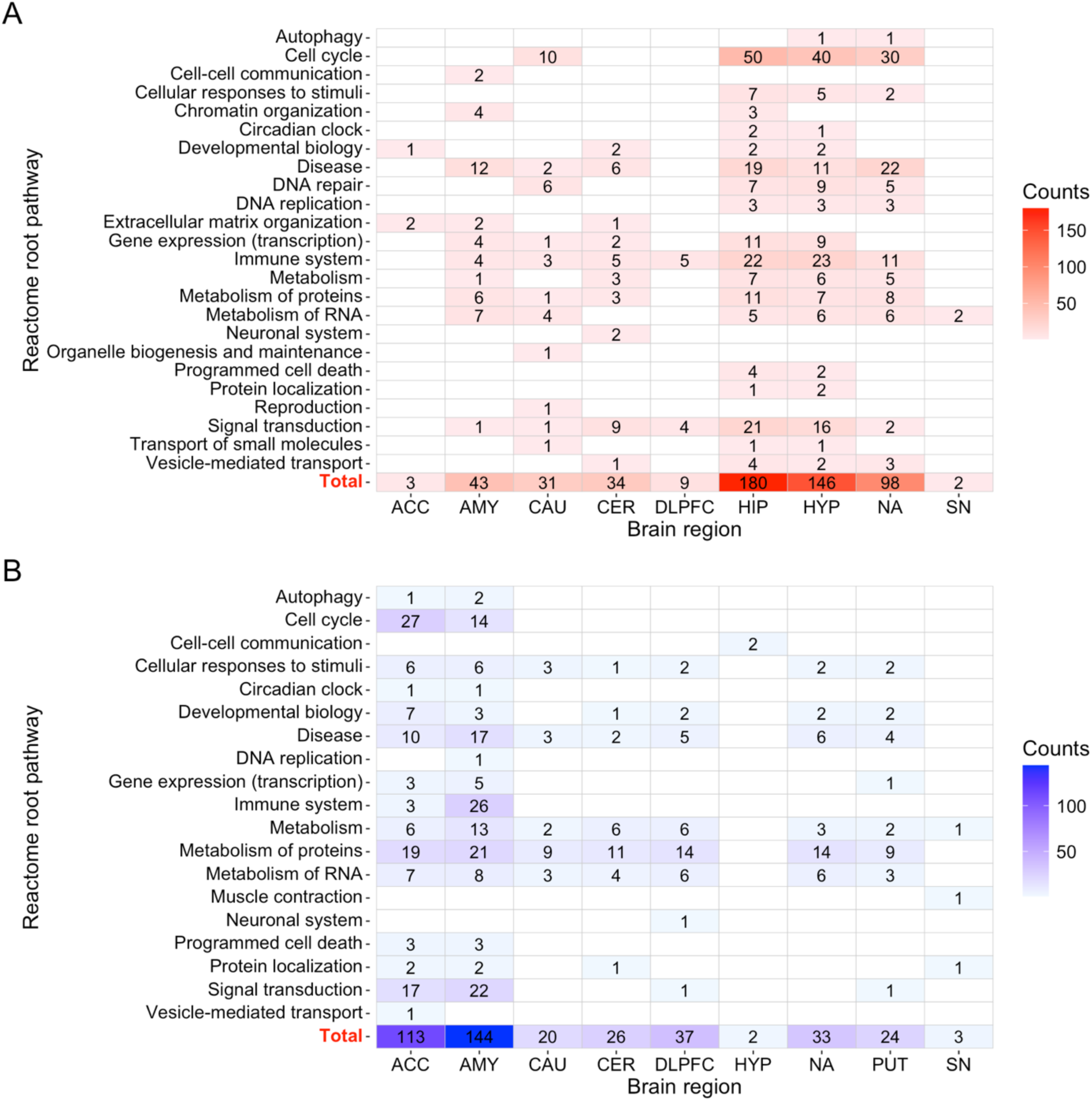
The number of over-represented Reactome sub-pathways within each root-pathway. This figure summarizes the Reactome pathways that were significantly over-represented by differentially expressed genes in the meta-analysis of each brain region. Panel A shows the Reactome pathways that were distinctly upregulated. Panel B shows the Reactome pathways that were distinctly downregulated. For each brain region, each over-represented pathway was traced down to a root-pathway term through the Reactome parent-child pathway hierarchy. In each brain region, the cells are labeled with the number of over-represented sub-pathways within the root-pathway. Cells with a larger number are highlighted with a darker-color shading. Abbreviations: ACC: anterior cingulate cortex; AMY: amygdala; CAU: caudate; CER: cerebellum; DLPFC: dorsolateral prefrontal cortex; HIP: hippocampus; HYP: hypothalamus; NA: nucleus accumbens; PUT: putamen; SN: substantia nigra.

We found two distinctly up-regulated Reactome pathways shared between those identified from imputed and directly measured DLPFC gene-expression profiles (Fisher’s exact test, Bonf.-p = 0.092; **Figure 4; Supplementary Table 7**), including RHOC GTPase cycle (R-HSA-9013106) and RAC2 GTPase cycle (R-HSA-9013404). One distinctly up-regulated Reactome pathway involved in RHOB GTPase cycle (R-HSA-9013026) was common in both imputed and directly measured cerebellum gene-expression profiles, but the overlap was not significant (Fisher’s exact test, Bonf.-p = 0.89). In addition, we found an overlap of 12 down-regulated Reactome pathways in DLPFC (Fisher’s exact test, Bonf.-p = 1.8 × 10⁻³) and 13 down-regulated Reactome pathways in cerebellum (Fisher’s exact test, Bonf.-p = 1.9 × 10⁻⁷), enriched for translation, mitochondrial translation, and related processes (**Figure 4; Supplementary Table 7**). Post-hoc analyses with single-cell expression data from *PsychENCODE* did not result in any significant enrichment of any cell-type-specific marker genes among the list of genes differentially expressed in AD found in the BrainGENIE-imputed brain-regional transcriptomes.

**Figure 4.**
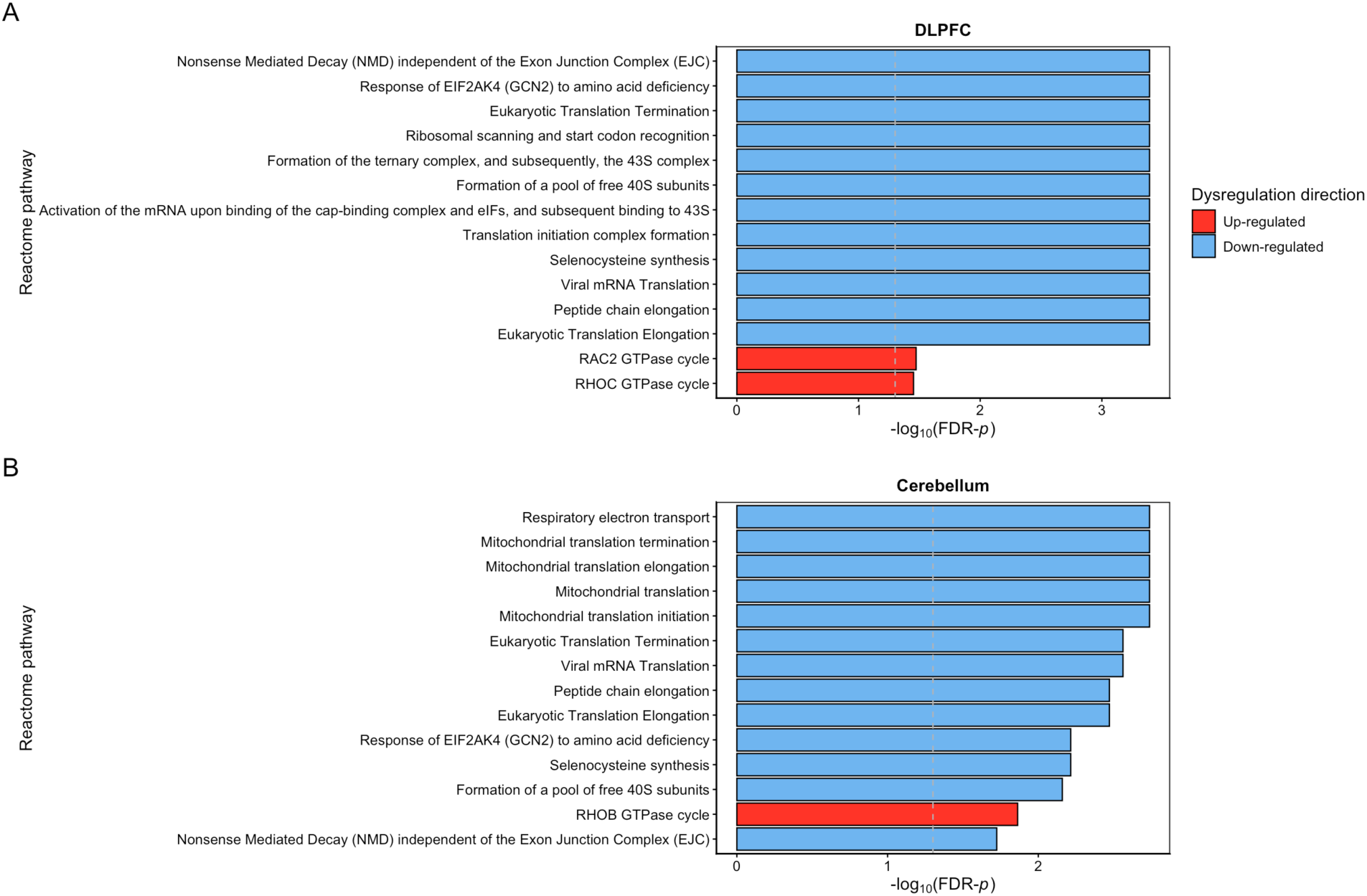
Reactome pathways shared between directly measured and imputed data. The figure shows the significantly over-represented Reactome pathways that were shared (Bonf-*p* < 0.05) between directly measured *postmortem* brain data and *BrainGENIE*-imputed data in DLPFC and cerebellum, respectively. Pathways in red color were distinctly up-regulated in AD. Pathways in blue color were distinctly down-regulated in AD. Abbreviations: Bonf-*p*: Bonferroni-corrected *p*-value; DLPFC: dorsolateral prefrontal cortex; AD: Alzheimer’s disease.

## Discussion

Transcriptome imputation offers promising opportunities to enhance insights into AD and complex brain disorders, opening possibilities for more targeted etiological exploration of healthy and diseased neural tissue. This study highlights the potential for *BrainGENIE* as a tool for analyzing AD pathophysiology. We discovered both reproducible and novel insights into the brain-regional gene expression changes associated with AD.

*BrainGENIE*-imputed brain gene-expression data showed significant congruence with data observed in postmortem brain, recapitulating changes among the most differentially expressed transcripts in addition to transcriptome-wide alterations associated with AD based on direct measurements of multiple distinct brain regions. The results indicate significant correlation between measured DLPFC and cerebellum data and data predicted by *BrainGENIE*, evidenced in both individual genes and shared pathways.

We did not find a significant correlation between directly measured and imputed gene-expression changes for AD in the hippocampus. We investigated potential reasons for this observation through multiple *post-hoc* analyses with the measured hippocampal data. The between-study correlations in the hippocampus measured datasets were not significantly different from those of the measured cerebellum and DLPFC datasets. As such, no marked irregularities were found in the datasets in each measured brain region, and dataset inconsistencies were ruled out as a bias source for the hippocampus measured data. We hypothesized that a potential source of biological variability in the measured hippocampal gene-expression data could stem from the profiling of different hippocampal substructures in the AD postmortem brain studies included in our meta-analysis. However, we were unable to test this hypothesis as the spatial-gradient classification model for the hippocampus failed to generalize on two independent postmortem brain AD studies (Blalock et al., 2004; Miller et al., 2008) where hippocampal substructure was known (accuracy <0.2). As a result, we could not exclude the possibility that sampling variability along the anterior-posterior axis or other intra-regional molecular gradients (*e.g.*, corresponding to hippocampal subfields) confounded the hippocampus-related results. So, the source of disparity between imputed and measured AD-associated gene-expression changes in the hippocampus remains unknown.

The weak correlation between *BrainGENIE*-imputed gene expression changes in AD and those found in measured postmortem hippocampus may be due to the extent of pathophysiological impacts on this region over the course of the disease. That is, the nature of gene expression may change with disease progression, so case-control comparisons may obscure stage-dependent effects that only manifest in subsets of cases. The current version of *BrainGENIE* captures a wide net of gene correlations in the heterogeneous non-AD samples available in GTEx V8. Future *BrainGENIE* iterations are being designed to widen the pool of predictable gene-expression profiles by adding datasets that include individuals at various stages of disease, with the goal of increasing precision of *BrainGENIE* output to a broader range of individuals (Hess et al., 2023).

Across imputed brain regions, the anterior cingulate cortex (ACC) and amygdala exhibited the highest number of AD-associated DEGs (705 and 687, respectively), followed by the caudate (52) and DLPFC. The rate of DEGs, which normalizes for the total number of genes analyzed, was highest in ACC (17.27%) and amygdala (11.89%). Notably, the substantia nigra showed no significant DEGs, suggesting either a lack of measurable transcriptional changes in this small region or a limitation of the imputation process. These results are congruent with studies highlighting the metabolic and morphological changes to the ACC and amygdala in AD (Al-Ani et al., 2023; Pardo et al., 2020).

Beyond validating some signals from *postmortem* transcriptomic studies of AD, *BrainGENIE* also has shed light on biological pathways previously not associated with AD pathology in the postmortem literature. Dysregulation in processes including chromatin organization, extracellular matrix remodeling, transport of small molecules, organelle maintenance, and DNA replication and repair was present in the imputed data. These processes are not considered hallmarks of AD in postmortem studies.

Genome-wide association studies of AD implicate microglial immune activation, extracellular-matrix remodeling, chromatin regulation (*e.g*., KAT8), mitochondrial oxidative-phosphorylation genes, ubiquitin–proteasome proteostasis, lipid/endocytic transport, DNA-repair/cell-cycle control and translation/ribosome pathways (Jansen et al., 2019; Kunkle et al., 2019; Bellenguez et al., 2022; Ghose et al., 2024; Paliwal et al., 2021; Simpson et al., 2016; Gentier & van Leeuwen, 2015). Our results show dysregulation in mRNA translation/ribosome biogenesis, DNA-replication-and-repair–linked cell-cycle re-entry, chromatin architecture, extracellular-matrix remodeling, organelle maintenance/autophagy–vesicle biogenesis and protein-synthesis–related cell-cycle processes that remain below genome-wide significance in the largest AD meta-GWAS to date (Jansen et al., 2019; Kunkle et al., 2019; Bellenguez et al., 2022). This suggests that *BrainGENIE* predictions of dysregulated gene expression align with some risk factors implicated in GWAS as well as potential changes downstream in the AD pathophysiological process.

Disruptions in chromatin organization may point to epigenetic changes. Extracellular matrix changes may reflect novel vascular or structural alterations in the brain. Impaired small-molecule transport and defective organelle biogenesis could hint at previously underappreciated aspects of cellular stress and metabolic failure. DNA replication and repair pathways showed alterations as well, raising intriguing questions about genomic instability in AD. The identification of these pathways suggests that *BrainGENIE* does not merely replicate findings from *postmortem* tissue, but instead expands the landscape of molecular abnormalities detectable in the living brain, offering new directions for understanding disease mechanisms and therapeutic targeting that could not have been found by studying the brain after death.

Beyond the identified changes in individual transcripts, our meta-analysis revealed significant dysregulation of multiple biological pathways, including those involved in protein homeostasis, immune activation, mitochondrial dysfunction, and neuronal signaling in the AD treatment group. Protein degradation pathways were significantly downregulated, supporting documented evidence for proteostasis dysregulation in AD (Thapa et al., 2024). Moreover, translation-related pathways were enriched among downregulated genes in the DLPFC and cerebellum, corroborating previous findings of impaired protein synthesis in AD, as evidenced in the amyloid hypothesis and the dysfunctional protein metabolism of APP by beta secretase which leads to insoluble amyloid formation (Hernández-Ortega et al., 2016). Studies show decreased ribosomal RNA (rRNA) integrity and reduced ribosomal proteins in AD, suggesting impaired ribosomal function (Patel et al., 2020; Hoozemans et al., 2009). eIF2α phosphorylation is increased in AD, leading to suppression of general protein synthesis while upregulating stress-related proteins such as ATF4, CHOP, and BACE1 (Koren et al., 2019).

Mitochondrial dysfunction was also suggested, with reduced expression of genes involved in oxidative phosphorylation and ATP production, consistent with prior studies demonstrating metabolic deficits in AD in multiple regions including the hippocampus (Ashleigh et al., 2023). Inefficient mitochondrial autophagy has been implicated both upstream and downstream of hallmark AD pathological processes of amyloidosis and tauopathy through increased oxidative stress (Mary et al., 2023). Moreover, downstream changes in mitochondrial fusion/fission as a result of amyloidosis have also been observed (Wang et al., 2008). This is also evident in familial AD, as presenilin, a protein family associated with familial AD and part of the gamma secretase protein involved in APP metabolism, is also present in mitochondrial-associated membranes and as such may provide evidence for the role of mitochondria in AD (Area-Gomez et al., 2009).

Immune system dysregulation was observed as well. This is in line with mounting evidence that neuroinflammation plays a central role in AD pathology, contributing to neuronal damage and disease progression (Sun et al., 2022; Taipa et al., 2019). Cell-cycle dysregulation was the top hit for both upregulated (in the hippocampus, hypothalamus, and nucleus accumbens) and downregulated (in the ACC) pathways. The literature supports the role of cell-cycle dysregulation leading to neuronal death in the two-hit AD pathological hypothesis, where cell-cycle reentry in response to oxidative stress (first hit) designed to lead to apoptosis dysfunctionally results in immortalized neurons (second hit) that provide the grounds for neurofibrillary tangles and plaques (Neve & McPhie, 2006; Moh et al., 2011).

## Conclusions

This paper demonstrates *BrainGENIE*’s effectiveness in imputing relevant regional brain transcripts in AD, recapitulating some of the known transcriptomic hallmarks of AD from studies of the postmortem brain, and revealing novel candidate transcripts and pathways in the brains of those living with AD that could not be identified via postmortem and blood-based gene expression analyses. Regional gene expression imputation offers the benefits of longitudinal monitoring and targeted and accurate regional representation that cannot be achieved via postmortem data and blood data respectively.

*BrainGENIE* provides a novel opportunity to investigate longitudinal changes in molecular markers of AD that can increase our understanding of changes in different stages of the disease process. Future iterations of *BrainGENIE*, incorporating training data from larger datasets with more paired blood-brain samples spanning a wider age range, or datasets enriched for brain disorders, may increase the imputation effectiveness for transcripts across various brain areas, enhance generalizability in both healthy and diseased states, and identify still more disease markers. Furthermore, reduced imputability of brain-regional transcripts, particularly in areas relevant to AD such as the hippocampus, could potentially serve as a biomarker for disease. Future directions for AD discovery include establishing better blood-based AD datasets that have multiple extraction timepoints and include clinical metrics such as Braak stage and MMSE scores. We encourage the field to consider the utility of expanding investment in gathering blood data for patients at various stages of disease as this can increase the power of imputation-based differential expression analyses and give us a higher resolution for the longitudinal pathological changes in AD. Ancillary investment in generating blood-based transcriptomic data has a clear strategic advantage to postmortem brain gene expression alone due to the increased accessibility, economic utility, and reduced confounding effect due to the absence of agonal factors. Such data can be utilized to investigate longitudinal DGE effects across different AD stages.

This project has established novel findings pertaining to pathological changes associated with AD and reconfirmed existing gene expression associations with AD through the usage of blood data and our state-of-the-art imputation algorithm *BrainGENIE*. Our results confirm many AD correlated pathways in the literature and further extend them through findings such as dysregulation in DNA and extracellular organization. As such, this study opens opportunities for investigation into plausible diagnostic and therapeutic agents associated with the reported pathological changes.

## Data Availability

All data produced in the present work are contained in the manuscript

